# Network for subclinical prognostication of COVID 19 Patients from data of thoracic roentgenogram: A feasible alternative screening technology

**DOI:** 10.1101/2020.09.07.20189852

**Authors:** Akash Bararia, Abhirup Ghosh, Chiranjit Bose, Debarati Bhar

## Abstract

**Background and Study Aim:** COVID 19 is the terminology driving people’s life in the year 2020 without a supportive globally high mortality rate. Coronavirus lead pandemic is a new found disease with no gold standard diagnostic and therapeutic guideline across the globe. Amidst this scenario our aim is to develop a prediction model that makes mass screening easy on par with reducing strain on hospitals diagnostic facility and doctors alike. For this prediction model, a neural network based on Chest X-ray images has been developed. Alongside the aim is also to generate a case record form that would include prediction model result along with few other subclinical factors for generating disease identification. Once found positive then only it will proceed to RT-PCR for final validation. The objective was to provide a cheap alternative to RT-PCR for mass screening and to reduced burden on diagnostic facility by keeping RT-PCR only for final confirmation.

**Methods:** Datasets of chest X-ray images gathered from across the globe has been used to test and train the network after proper dataset curing and augmentation.

**Results:** The final neural network-based prediction model showed an accuracy of 81% with sensitivity of 82% and specificity of 90%. The AUC score obtained is 93.7%.

**Discussion and Conclusion:** The above results based on the existing datasets showcase our model capability to successfully distinguish patients based on Chest X-ray (a non-invasive tool) and along with the designed case record form it can significantly contribute in increasing hospitals monitoring and health care capability.

## I. Introduction

Towards the completion of a semi-annual tenure beginning from the first reported case of COVID-19 on 29^th^ January, 2020 in India, there has been a substantial rise in the number of new cases and associated death rates across the country. Amongst 21 countries, a total of 9996 cases has been reported as on 30^th^ January, 2020^[1]^. COVID-19 is an extremely contagious disease capable of producing a global threat to health based on benchmark criteria such as prevention, diagnosis and treatment^[2]^. According to WHO, 16,341,920 confirmed cases has been reported as on 30^th^ July, 2020 out of which deaths occurred in 650,805 cases. In India alone, 1,531,669 confirmed infected cases have been reported out of which 34,193 died as on 30^th^ July, 2020.

The SARS-COV2 viral disease has similarities with influenza in the early stages, which During an approach to develop a mass screening makes it very difficult to diagnose. On the contrary, late diagnosis can lead to the death of the subject as well as spread of infection. During an approach to develop a mass screening procedure, a lot was accessed about the quantum of errors that might be there in the current molecular as well as the serological procedures like RT-PCR and antibody tests. Since none of the above tests are accurate and in the scenario of the non-availability of a gold-standard procedure, the choice of test is based on its sensitivity and specificity. Country-wide RT-PCR based approach to detect viral RNA is currently being used^[3, 4]^.

Owing to the rise in COVID-19 cases across the globe, systemic utilization of healthcare resources is of crucial importance. Machine learning technologies can be a significant strategy in this situation for proper futuristic prediction that can help the administrators to plan accordingly in terms of health care resources and its associated support mechanisms. As the onset of disease is primarily in the lung of the infected person, so images of the lung could give us a quicker and better predictive approach. But to make it quicker, we need to use artificial intelligence and machine learning as a front-line analytical tool of the images^[5–7]^.

Our main goal in this work is to develop a neural network-based prediction model that is capable of distinguishing the COVID-19 viral infected patients from normal people as well as from bacterial pneumonia based on the images of thoracic roentgenogram. The model takes an input of the X-ray image and then analyses it to state the outcome. The benefit of this procedure is based on user flexibility and associated facts: X-ray test is cheap, very fast image development time, availability of the instrument in most clinics and hospitals across the country. The additional benefits include scanning of the x-ray plate and uploading it into the network doesn’t always require skilled manpower or medical personnel, hence it is effective in mass screening and helps to reduce burden on skilled hospital labour and prevents overcrowding of hospitals. Associatively a Case record form (in Supplementary file) is developed that has certain essential input spaces along with the network predicted result which needs to be filled and can be done by anyone without prior training. Based on collective outcome of the rest of inputs and network result, an initial screening is possible and then, if needed, the patient can be sent to doctors based on the government and hospital workflow guidelines. A schematic representation of the combined model is stated in figure 1.

**Figure 1:**
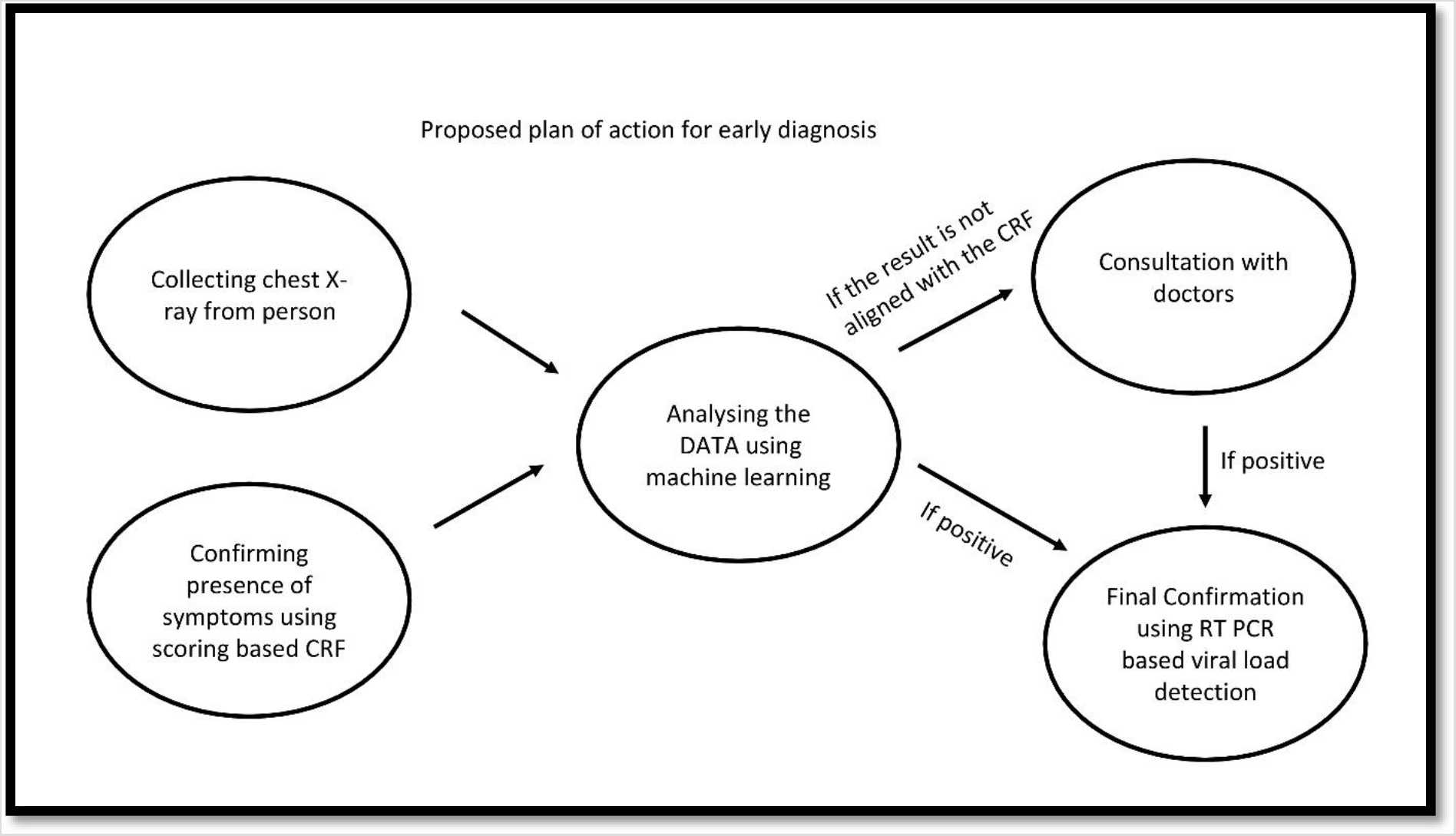
Prognostication model developed by the successive use of the neural network-based prediction model and the associative case record form developed.

## II. Materials and Methods

### II.I Data source/Dataset Development and structuring

During the course of this study, all data gathered is in the form of jpeg image files of thoracic roentgenograms. These files have been acquired from different sources and have different sizes. The main two main sources of Chest X-ray images were taken from GitHub (https://github.com/ieee8023/covid-chestxray-dataset) and Kaggle (https://www.kaggle.com/paultimothymooney/chest-xray-pneumonia). These two datasets have been fused and their data used to create our final dataset comprising of equal proportions of COVID-19, bacterial, other viral and normal chest X-ray images^[8, 9]^. We have also assumed that COVID-19 cases and viral pneumonia have similar symptoms and so these images are merged into a single feature. After acquiring the images, we have created a pandas data-frame of the image path and its corresponding label. Thus, our final dataset is a pandas data frame containing 3840 images, which are equally distributed.

### II.II Data cleaning and Data Augmentation

Preliminary data cleaning steps involved validating the image’s readability and conversion. Since most of the images obtained were already pre-cleaned and verified, this step ran relatively fast and only a small percentage, about 0.1% of the images were found to be corrupted and hence were rejected from the study. Since the dataset has multiple images with different image sizes, some amount of pre-processing had to be performed. The pre-processing step firstly converted these images to a single channel gray-scale image.

After this step, slight image enhancement techniques were done by passing it through a sharpening filter. The final step was to convert the image to a 200*200 matrix and normalising the values.

Also, in order to satisfy limited memory usage during the training phase of the model, a suitable data generator was created which could generate random images of batch size 10 and feed it to the network. This data generator was used for training as well as in the serving phase, and all pre-processing steps used during training were also applied during the production phase. The protocol structuring for the experiment was done in a systematic manner as indicated in figure 2.

**Figure 2:**
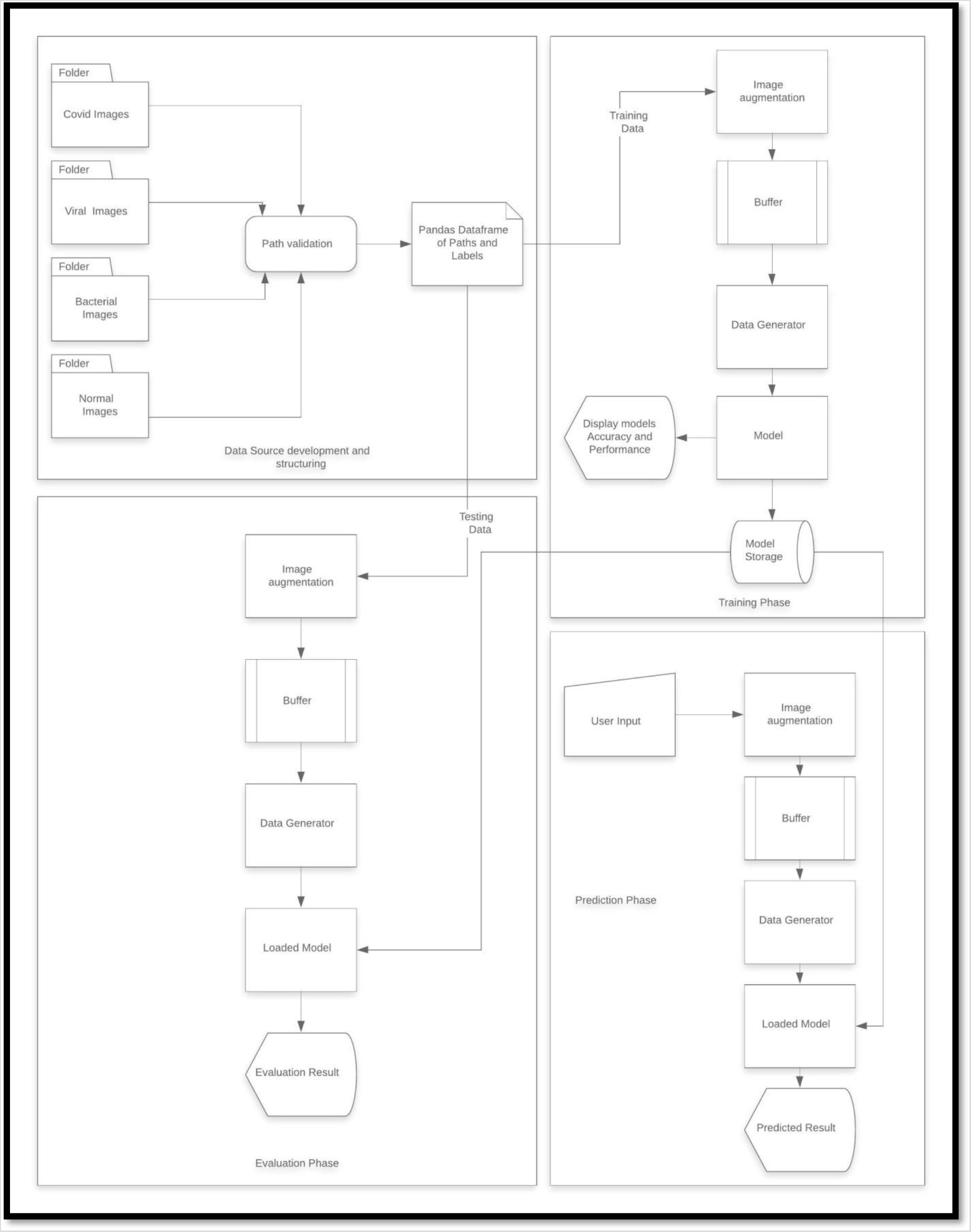
Architecture of the system used to develop the neural network-based prediction model.

### II.III Computational setting

The training of the model was computationally expensive, primarily because it involved convolutional neural networks, being trained on about 1000 images. This required high memory and processing capabilities and so conventional laptops or PC’s could not satisfy this. Hence majority of the training was carried out on a python notebook hosted on Google Cloud platform. The underlying system configuration was 8 CPU’s, 32 GB of RAM and 1 TB of hard disk space. The models were written in Keras using a tensor flow backend.

Owing to the fact that Google Cloud platform has out-of-the-box support for tensor flow, to ensure production level robustness and ease of deployment, it was chosen and all the training was done on this platform. This also gave us the capability to deploy the model in future in a scalable manner without the hassle of setting up servers and configurations.

### II.IV Statistical analysis

Kolmogorov-Smirnov test was used to determine the distribution of the subjects. Then for the group of subjects having normal distribution, unpaired t-test used. If one or both groups didn’t have normally distributed data, Mann-Whitney test was used for comparison. All statistical analysis was performed using GraphPad Prism version 8.0.2.

## III. Results

### III.I FEATURE SELECTION

There were not many feature selection steps involved in the study and hence all of the feature selection steps have been omitted or carried out in the pre-processing steps.

### III.II NETWORK ARCHITECTURE AND OPTIMIZATION

The input layer of the network consists of a 2D convolutional network with 32 filters followed by a dropout layer. This is followed by another 2D convolutional layer with 64 filters and another dropout layer. This is again followed by 3 layers of a dense network, each with 64, 32 and 16 cells respectively. The layers are activated using a RELU activation function. The output layer consists of a soft-max layer with 3 output nodes. Categorical Cross Entropy is selected as the loss function and the layers are trained using an Adam optimizer. The final model consists of 157,373,475 trainable parameters. The output of the model is a probability distribution spread across 3 class labels.

### III.III TRAINING

The model was trained on a batch size of 10 images each of 200 * 200 in dimension. Furthermore, the model was trained for 20 epochs and the dataset was shuffled after the end of each epoch. The total number of samples trained in each epoch was 2688. The total training time of the model was 1hour 15 minutes on the hardware. This time could be further reduced by training on a lower number of samples or increasing the computational power of the system. At the end of the training, the model was saved as a .pb file in the local file system which could be later used to recreate the model. After multiple iterations, the best performing model was chosen and was used for final evaluation study.

### III.IV Prediction accuracy

The model gave an overall accuracy of 82% among the 3 different classes. This can be further expanded as the following scenarios: -

- The model provided 93.2 % accuracy when trying to detect whether a person is normal or not i.e. it is able to differentiate a normal person from an infected person with very high accuracy.
- The model is able to detect with 74.9% accuracy that a person has Bacterial pneumonia whereas in the rest cases it detects the person as having Viral pneumonia.
- The model is able to detect with 76.5% accuracy that the person has viral pneumonia whereas in the rest 23.4% cases it detects that the person has bacterial pneumonia.

| Actual/Predicted | Bacterial | Normal | Viral/COVID-19 |
| --- | --- | --- | --- |
| Bacterial | 251 | 9 | 75 |
| Normal | 5 | 302 | 17 |
| Viral/COVID-19 | 68 | 12 | 261 |

Furthermore, other performance parameters of the model are as follows: -

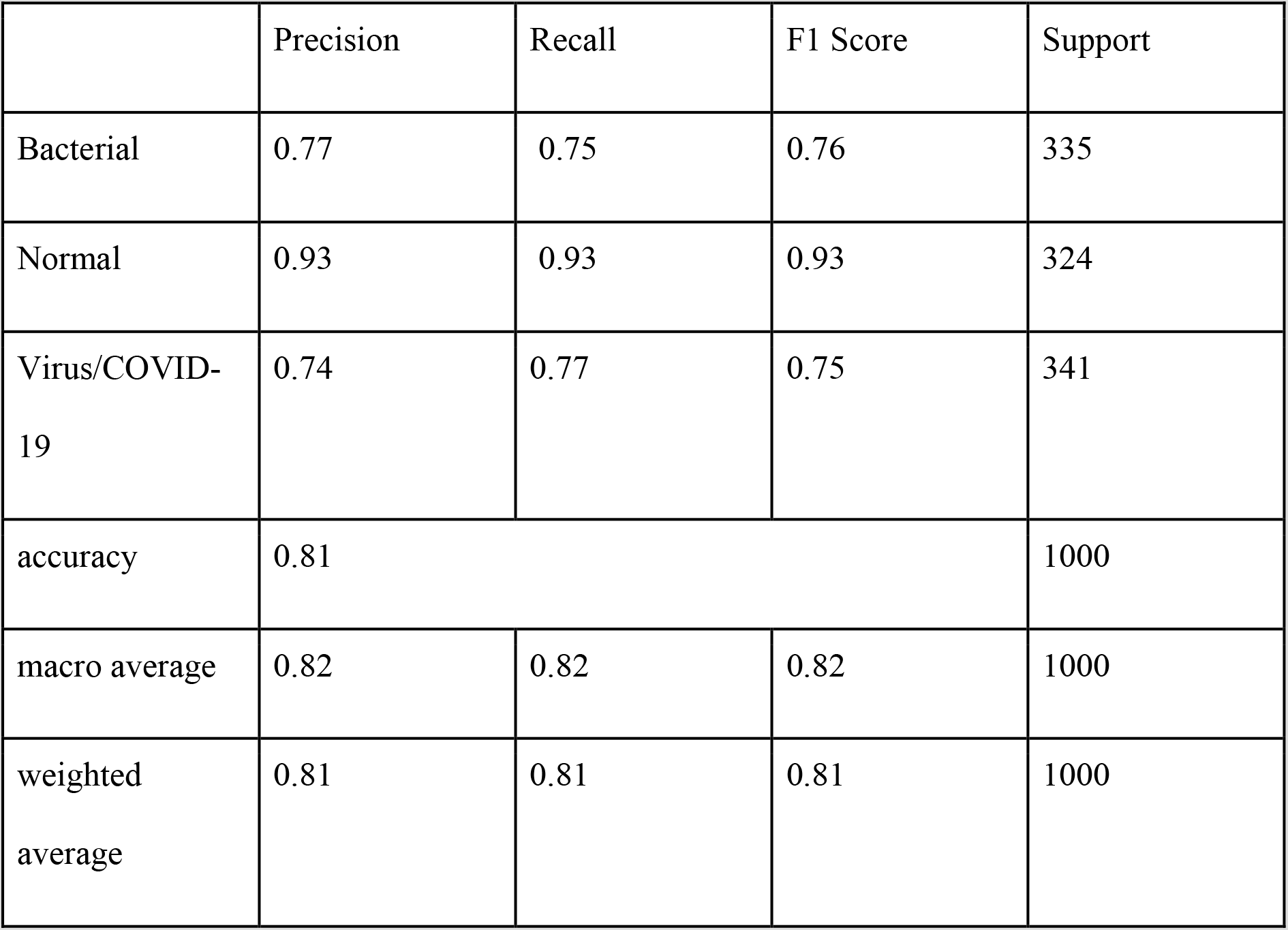

The AUC score for the above data is 93.7%. Final accuracy of the model as tested on 1000 samples was found to be 81% and the sensitivity is 82% with a specificity of 90%. With more and better-quality data, the accuracy can be further increased. The performance metrics of the data was obtained using the confusion matrix with the help of Scikit Learn library.

### III.V Statistical analysis

COVID-19 is a respiratory pathogen caused by SAR-COV2 virus which has a probable zoonotic origin. It mainly enters the body via the upper aero-digestive tract, and produces a deadly attack on the lungs of the infected person with severe disease. Subjects predominantly have respiratory distress or even worse being ‘happy hypoxia’, i.e. a state of oxygen desaturation without significant shortness of breath and hence leads to a crucial delay in seeking medical help, which might result in fatality. Mainstay of supportive therapy is oxygen support and/or ventilation. We assumed that oxygen saturation may play a crucial role in the survival of these patients. But from the datasets we saw that there was no significant difference in the oxygen saturation level of the subjects who died of SARS-COV2 viral illness vis-a-vis those who recovered from the disease as shown in Figure 3. Hence, we may say that although oxygen saturation is an important clinical marker of the disease, it probably doesn’t play a big role in predicting mortality.

**Figure 3:**
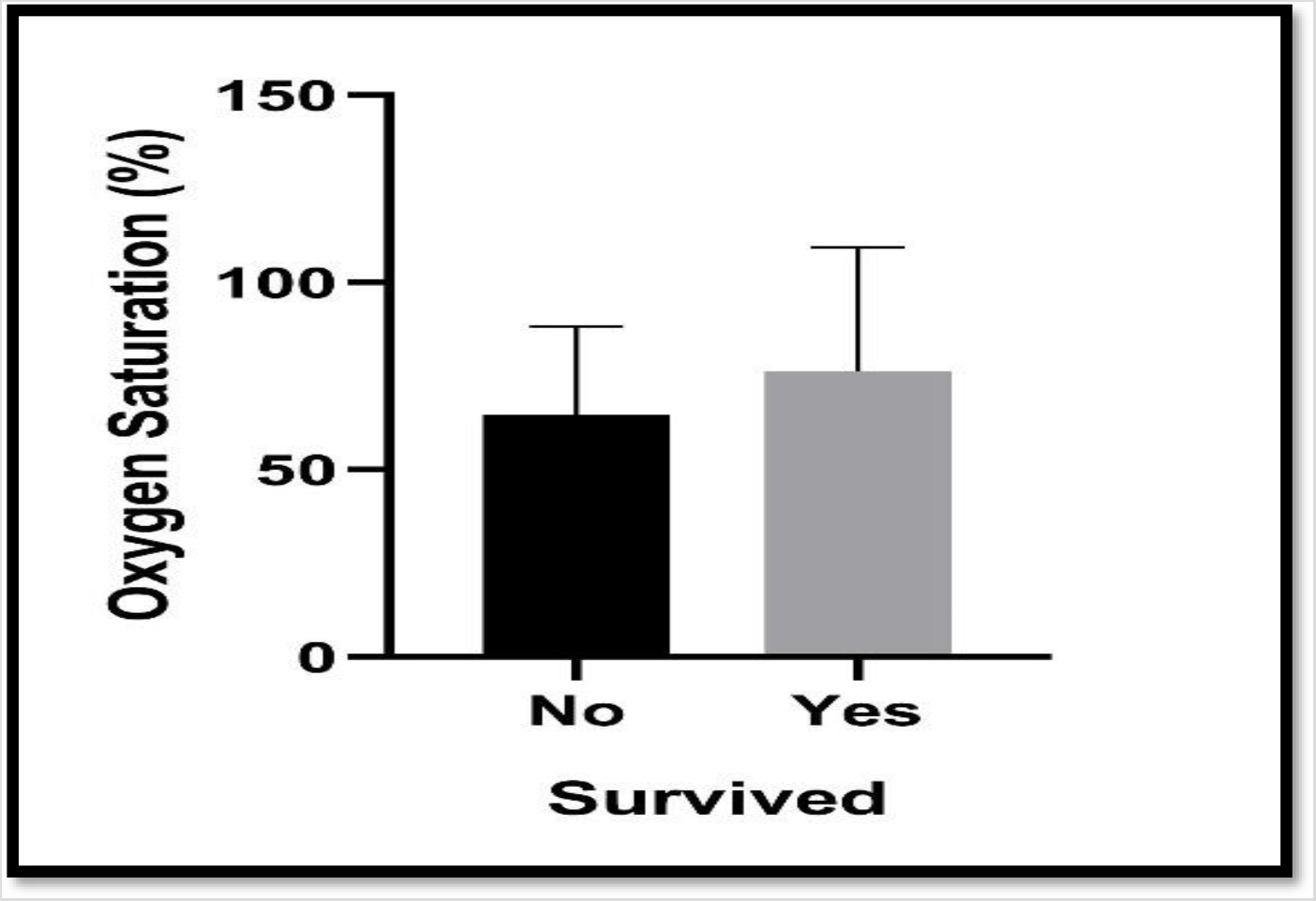
Bar Chart representing that there is no statistically significant difference observed in SpO2 level between the subjects that died due to COVID –19 and those who survived after being infected.

## IV. Discussion and Conclusion

The first documented case of corona virus appeared in 1960 followed by the death of about a thousand patients in 2003, and finally a pandemic led by a different strain of corona virus in 2020. Irrespective of such a long history of affliction of the human race, scientists have been unable to produce an officially available vaccine against this deadly air-borne virus^[10]^.

Evidence of pathogenicity is directed by the fact that it can spread via air-borne droplets and via asymptomatic patients across all ages which makes every age group susceptible. Even patients can spread this virus after immediate recovery^[11]^. Hence, under the prevailing circumstances, there is an urgent need to keep health-care officials safe from nosocomial infection of COVID-19. This can be only done by early diagnosis followed by isolation, and decrease of the patient burden on hospitals and associated intensive care units (ICU). This scenario might be critically dealt with by implementation of machine learning based methodology like a dynamic neural network based on thoracic roentgenogram^[12]^. Nevertheless, the only drawback comes from the dataset development part as developing a dataset during a pandemic is troublesome but very crucial for strategic planning in healthcare organisations^[5–7]^. However, we predict that our model along with case record form (CRF) can significantly contribute in quicker and cheaper way, for mass screening of patients (non-invasively) and also help in managing and streamlining health care centres and its associative monitoring

## Data Availability

The datasets used and/or analyzed during the current study are available from the corresponding author on reasonable request

https://github.com/ieee8023/covid-chestxray-dataset

https://www.kaggle.com/paultimothymooney/chest-xray-pneumonia

## Credit authorship contribution statement

**Akash Bararia:** Conceptualization, Data sorting and mining, original draft.

**Abhirup Ghosh:** Data curation and augmentation, Computational analysis, Writing – original draft.

**Chiranjit Bose: Statistical analysis**, Writing – original draft, Editing, Dataset curation.

**Debarati Bhar:** Supervision, Clinical Correlation and analysis, Draft proofreading.

## Ethical Approval

None

## Acknowledgement

We would like to thank all the patient and normal individuals whose data has been incorporated into the datasets. We would also like to thank Prof. Dr. Satinath Mukhopadhyay, Professor at the Institute of Post Graduate Medical Education and Research (IPGME&R) in Kolkata for his motivational support. We would like to thank The Department of Science & Technology for financially supporting Mr. Chiranjit Bose via DSTINSPIRE FELLOWSHIP (Inspire fellow registration number: IF180004). Alongside We would also like to thank our respective employer’s. We would also like to thank Joseph Paul Cohen, Director of the Institute for Reproducible Research, for GitHub dataset and Paul Mooney, Developer Advocate at Kaggle for the Kaggle dataset development. Last but not least we would also like to thank all doctors and health support teams working across globe to fight against coronavirus pandemic.

